# Entrenching social norms in Community-led total sanitation for sustainability of open defecation free status: A survey of Suna West Sub-County, Migori County, Kenya

**DOI:** 10.1101/2022.12.05.22283135

**Authors:** Naomi R. Aluoch, Collins O. Asweto, Patrick O. Onyango

## Abstract

**Background:** Community-led total sanitation (CLTS) has been used to stir sanitation-related behaviour change and attain open defecation free (ODF) status. CLTS interventions suffer high rates of reversion such that their gains are unsustainable in most contexts including Suna West sub-County, Kenya.

**Objective:** This study aimed at determining the role of sanitation hygiene practices and social norms on open defecation free status in Suna West Sub County.

**Methodology:** Survey study design was employed using questionnaire and observation checklist to collect data from 384 households.

**Results:** Results revealed that 66.1% households had partially reverted to non-ODF status. The sanitation-hygiene practices associated with maintenance of ODF includes: use of treated water (OR=3.17; CI=1.20-8.40; *p*=0.020), use of elevated racks (OR=2.17; CI=1.08-4.37; *p*=0.030), regularly clean latrines (OR=4.88; CI=1.12-21.37; *p*=0.035), pouring of ash over the pit of the latrine (OR=4.25; CI=4.20-8.87; *p*<0.001) and use of dug out pits for waste disposal (OR=4.51; CI=2.09-9.78; *p*<0.001). On social norms, the study found that laws/penalties (OR=0.31; CI=0.21-0.48; *p*<0.001), need to improve things in the family (OR=0.50; CI=0.28-0.92; *P*=0.025), and rewards/incentives (OR=0.21; CI=0.13-0.33; *p*<0.001) would reduce odds of being ODF. Moreover, odds of being ODF was less likely for households with perception that; construction/maintenance materials were expensive (OR=0.52; CI=0.33-0.80; *p*=0.003), most people don’t have a latrine (OR=0.40; CI=0.25-0.64; p<0.001) and it is okay to defecate in bushes/rivers/dams (OR=0.31; CI=0.19-0.51; p<0.001).

**Conclusion:** This study findings provides evidence of ODF status reversion in previously certified villages. However, household with retained ODF status was enhanced by several sanitation hygiene practices. Interestingly, households that displayed social norms were less likely to be ODF. This reveals that the CLTS process failed to instil social norms around proper sanitation to inspire community collective action thus little influence on sustainable behaviour change. The findings of this study therefore highlight the need to enhance good hygiene sanitation practices, while instilling social norms to inspire community collective action.

## INTRODUCTION

According to the WHO, roughly 842,000 lives are lost in low- and middle-income countries annually as a consequence of inadequate water, hygiene and sanitation (WHO, 2018). Poor sanitation is connected to infections such as diarrhoeal diseases, nematode infections and environmental enteropathy (EE) (UNICEF., 2015c). In Kenya diarrhoea claims the lives of roughly 3,100 children annually and trachoma, schistosomiasis are health problems linked to poor sanitation (Mutambo, 2016). In part the burden of these diseases is attributed to open defecation that exposes a large part of the population to sanitation-related diseases (Njuguna, & Muruka, 2017).

It is in light of such negative impacts of poor sanitation that the Government of Kenya adopted Community-led Total Sanitation (CLTS) as a strategy to improve sanitation. Community-led Total Sanitation was introduced by Plan International Kenya in 2007 and was approved by the then Ministry of Public Health and Sanitation (MOPHS) as a key framework for promoting hygiene and sanitation at the household level. In 2011, MOPHS established CLTS as the national strategy for ensuring rural sanitation and set a national target to reduce open defecation (Crocker, Saywell, & Bartram, 2017).

The results of a study on the sustainability of ODF status in Kenya conducted by UNICEF (2015) revealed that the sustainability of ODF achievements remained a major concern with over 70% of villages that had received partial or full ODF status reverting to non-ODF status. Among the factors that demotivate community members from using a latrine after becoming ODF relates to physical aspects of the latrine (such as lack of privacy and fear of the latrine collapsing) and sharing a latrine with other people (Singh, & Balfour, 2014). In addition, slippage from ODF status has also been linked to collapse or poor structural integrity of latrines as well as unsustainable behaviour change following sanitation-related interventions (UNICEF, 2014). This study investigated association between ODF status with sanitation-hygiene practices and social norms in a previously ODF certified region, Suba West Sub-County, western Kenya.

## MATERIALS AND METHODS

### Study site

The study was done in Suna West Sub-County in Migori County which has a population of 117,539 with a density of 406 persons per km^2^. The sub-county, one among eight others, has four wards and is bordered by Kuria West sub-county to the south-east, Nyatike sub-county to the west, Suna East Sub-County to the north-eastern side and Tanzania to the south-west.

### Study Design

A cross-sectional study design was used across two wards that were purposively chosen for having attained ODF status in all the villages at least one year to the study. The unit of analysis was the household with the targeted participants being the household heads.

### Data Collection Tools

Validated structured questionnaire and observation checklist were used in this study. Observation checklist was used to collect information to corroborate or refute claims made by respondents in questionnaires.

### Study Variables

In order to determine the ODF status, re-verification was done using the verification tool focussing on the non-negotiable indicators used during the sub-county verification and third-party certification. In brief, the non-negotiable indicators that the tool focused on were no exposed faecal matter, access to latrine (individual or shared), privacy on superstructure, squat hole cover and hand washing facility near the latrine. In each household, we focused on the 5 non-negotiables which had to be in every household for it to be assessed as ODF.

### Dependent Variables

Dependent variables were measured as follows: access to a latrine - availability of Individual latrine, shared latrine/neighbours; privacy - availability of door or some form of barricade provided for each superstructure; Squat hole cover - provided for every squat hole and in use; hand washing facility - availability of tap/leaky tin near latrine with water inside, soap/ash available; no exposed faeces - no visible faeces within the surrounding of the home.

### Independent Variables

For independent variables, the frequency of variables such as treating water (boiling or use of chemicals), covering food using lid over cooking pots when cooking and during storage, using elevated racks to hold utensils off the ground while drying, regular cleaning of latrine, application of ash around & the squat hole of latrine, and using dugout pit for waste disposal were measured using Likert scale: always, most of the time, sometimes, rarely, not at all and scores pooled into two - Yes (always, most of the time, sometimes) and no (rarely, not at all).

While variables such as Care of the family - Empirical & normative expectation regarding health of the family, Shame/disgust/fear/pride Regrettable occurrence/ unpleasant emotion that cause a feeling of resolution, Cultural/social/religious beliefs - Person’s belief alignment as pertaining culture, society and religion, Laws/penalties - Rules within a given set up and punishment imposed for breaking the set rules, Need to improve things in the family - Empirical & normative expression of obligation to make things better, Follow ups and support - The subsequent actions following CLTS and material assistance for the same, Rewards/incentives - Some form of payment given in recognition of work done or to stimulate greater output, and Peer pressure - The empirical &normative expectation regarding consistent latrine use were measured using Likert scale: strongly disagree, disagree, neutral, agree, strongly agree and scores pooled into two; Yes (agree and strongly agree), No (strongly disagree, disagree, neutral).

### Statistical Analysis

Summation for the observed non-negotiable indicators was done to determine the ODF status. Wilcoxon signed-rank test was used to determine if there was a significant difference in ODF status as at the time of the study and verification. Chi-square test of independent was used to determine association between sanitation and hygiene practices, social norms and ODF status and binary logistic regression was done to determine the relationship between sanitation hygiene practices, social norms and ODF status.

### Ethical considerations

Ethical approval for the study was granted by the Maseno University Ethics Review Committee; Ref: MSU/DRPI/MUERC/00821/99.

### Informed consent

Informed consent was gotten from the participants. They were also informed that taking part in the study was out of free will and that they were free to withdraw from the study at any time.

## RESULTS

### Socio-demographic characteristics of study participants

Of the 384 participants, 62.8% were females, 73.4% were aged 25-59 years, while 58.6% and 31.8% had primary education and secondary education respectively. On socio-economic status, three-quarter (75.3%) of households had Ksh. 0-5,000 monthly income. A half (53.1%) of the households had 0-5 years old child and 27.1% had at least one member with a disability or chronic illness. Socio-demographic characteristics of the study participants are summarized in Table 1.

**Table 1:** Socio-demographic characteristics of study participants from Suna West Sub-County, Kenya.

|  | **Variable** | **Frequency**<br>**(n)** | **Percentage**<br>**(%)** |
| --- | --- | --- | --- |
| **Gender** | Male | 143 | 37.2 |
|  | Female | 241 | 62.8 |
| **Respondents' Age** | 18-24 years | 58 | 15.1 |
|  | 25-59 years | 282 | 73.4 |
|  | 60 and above | 44 | 11.5 |
| **Level of Education** | No education | 26 | 6.8 |
|  | Primary education | 225 | 58.6 |
|  | Secondary education | 122 | 31.8 |
|  | Tertiary education | 11 | 2.9 |
| **Level of Income** | 0-5,000 | 289 | 75.3 |
|  | 5,001-10,000 | 72 | 18.8 |
|  | 10,001-20,000 | 6 | 1.6 |
|  | 20,001-30,000 | 3 | 0.8 |
|  | 30,001-40,000 | 6 | 1.6 |
|  | 40,000 and over | 8 | 2.1 |
| **Household composition** | With persons 0-5 years. | 204 | 53.1 |
|  | With persons 6-12 years. | 278 | 72.4 |
|  | With persons 13-24 years. | 299 | 77.9 |
|  | With persons 25-59 years. | 354 | 92.2 |
|  | With persons above 60 years. | 72 | 18.8 |
|  | Persons with disability or chronic illness | 104 | 27.1 |

### Open Defecation Free Status

On ODF status, only 33.9 % (n=130) were found to be ODF one year after certification. When the indicators were analysed singly, it was observed that access to latrine and no exposed faeces were at 100%; with 95.3% (n=366) owning individual latrines while the remaining 4.7% (n=18) reporting to use shared latrines, Table 2.

**Table 2:** Results on ODF indicators.

| Indicator | Median percentage (%) | P value | No. of villages reporting 100% |
| --- | --- | --- | --- |
| Access to latrine | 100 | 1.0 | 13 (100%) |
| Squat hole cover present | 63 | 0.002 | 0 (0%) |
| Privacy | 82.4 | 0.002 | 0 (0%) |
| Hand washing facility | 82.4 | 0.004 | 2 (15%) |
| No exposed faeces | 100 | 0.056 | 8 (61.5%) |

### Association between sanitation and hygiene practices and open defecation free status

This study found association between sanitation and hygiene practices and ODF status. The results showed that households that treated water, used elevated racks, regularly cleaned their latrines, poured ash over the pit of the latrine and used dug out pits for waste disposal were more likely to be ODF; Table 3.

**Table 3:** Association between sanitation and hygiene practices and open defecation free status in Suna West Sub-County, Kenya.

| Characteristic | Use | ODF (n) | NOT ODF (n) | P Value | OR (95%CI) | P value |
| --- | --- | --- | --- | --- | --- | --- |
| Treating water | Yes | 123 | 225 | **0.015** | 3.17(1.20 - 8.40) | **0.020** |
|  | No | 5 | 29 |  |  |  |
| Covering food | Yes | 128 | 250 | **0.305** |  |  |
|  | No | 0 | 4 |  |  |  |
| Using elevated racks | Yes | 117 | 211 | **0.027** | 2.17(1.08 - 4.37) | **0.030** |
|  | No | 11 | 43 |  |  |  |
| Regular cleaning of latrine | Yes | 128 | 236 | **0.026** | 4.88(1.12 - 21.37) | **0.035** |
|  | No | 2 | 18 |  |  |  |
| Pouring of ash | Yes | 121 | 193 | **<0.001** | 4.25(2.04 - 8.87) | **<0.001** |
|  | No | 9 | 61 |  |  |  |
| Dug out pit for waste disposal | Yes | 122 | 196 | **<0.001** | 4.51(2.09 - 9.78) | **<0.001** |
|  | No | 8 | 58 |  |  |  |

### Association between social norms and open defecation free status

This study found association in a number of the social norms and ODF status. The social norm variables found to be associated with ODF include Subjection to laws and penalties, need to improve things, follow-ups and support, construction/maintenance expensive, majority ashamed for not having latrine, and okay to defecate in rivers/bushes/dams as shown in Table 5.

**Table 5:**
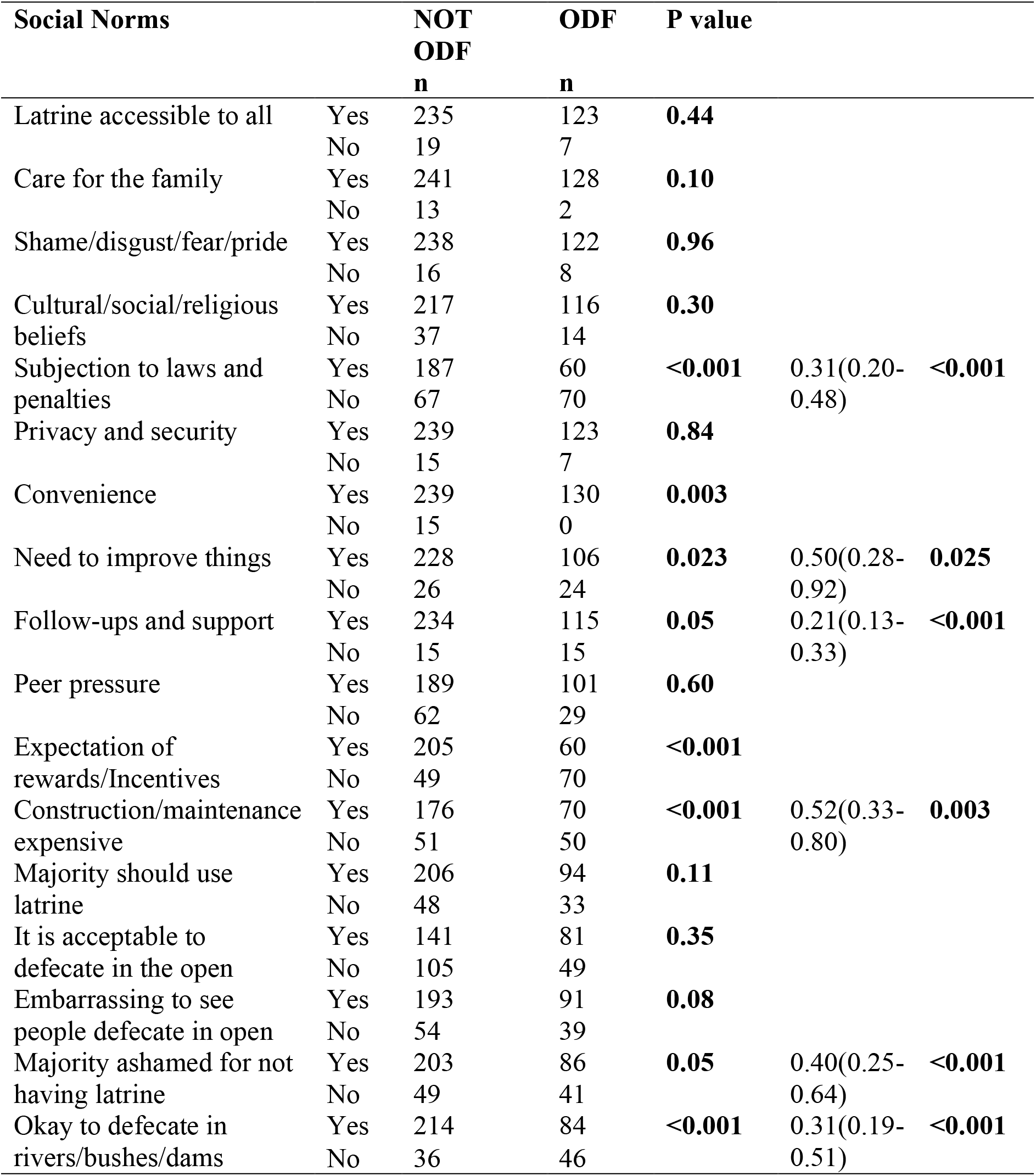
Association between social norms and open defecation free status Suna West Sub-County, Kenya.

## DISCUSSION

### Open defecation status

This study found 66.1% reversion one-year post-ODF in a previously ODF-certified region. Similarly, earlier study had shown 70% of villages reverting back to open defecation three years after certification among seven sub-counties featured in the study (UNICEF, 2015). However, low reversion rates have been observed (8%) in Ethiopia and Ghana after one year of CLTS implementation, and (14.5%) in Indonesia after two years of ODF certification (Crocker, Saywell, & Bartram, 2017; Odagiri, et al., 2017). In these studies, latrine presence - latrine status and usage – was used as the measure for sustainability, the possible reasons for recording lower reversion rate. Nevertheless, reversion has been found to be common in villages within sub-Saharan Africa where it has been associated with several factors (UNICEF et al., 2013; Mukherjee, 2012). Moreover, sustainability of ODF achievements has been previously found to be a major challenge in Kenyan communities.

In a study by Tyndale-Boscoe et al. (2013) in Uganda, Kenya, Ethiopia, and Sierra Leone two years after CLTS, a 13% reversion was reported when latrine presence was used to measure sustainability. However, the reversion rate would have drastically increased to 92% had the study used the 5 indicators used during the initial verification process which included functional latrine, means of keeping flies away (water seal or squat hole cover), absence of faecal matter, presence of hand washing facility with soap/ash and evidence of latrine use in the re-verification(Tyndale-Boscoe et al., 2013); a much higher reversion rate like the 66.1% observed in the present study when all the indicators are used to measure sustainability.

While governments and most organisations have been very successful in getting households to build and retain latrines, less success has been achieved in improving sanitation behaviour change which is the major aim of CLTS (Tyndale-Boscoe et al., 2013). Overall, the findings of this study suggest that there is need to harmonise or standardize, across studies, indicators that define ODF status. Furthermore, although the protocol is very clear on the non-negotiable indicators, there is need to re-look at their role in defining ODF status. Doing so will help in defining concepts up front in developing any kind of monitoring tool of post-ODF status.

### Association between sanitation hygiene practices and open defecation status

We found a significant association between non-negotiable sanitation hygiene practices and ODF status and demonstrated that households that complied with the sanitation hygiene practices were more likely to be ODF. Odagiri et al. (2017) noted in their study that participants from better performing villages on ODF outcomes reported that messages around sanitation promotion and good hygiene had been constantly promoted through mosques and churches. Further, local groups carried out monitoring after CLTS implementation in an effort to promote hand washing with soap, treating of drinking water, proper food handling, solid and liquid waste management by households.

Maintenance of ODF status in households that poured ash in the pit latrines is not surprising because pouring of ash in the pit latrines manages smell from latrines and therefore encourages consistent latrine use by all members of the household. This finding resonating with previous study by Mukherjee (2016) which reported that smelly and unimproved latrines turned people back to open defecation and in Ethiopia, latrine usage by women was tampered with negatively as a result of perceptions around latrine cleanliness and smell inside (Odagiri, et al., 2017). While the present study found that 21% of households presented no evidence of the use of a hand-washing facility and 46% households did not wash their hands with soap and water always after using a latrine, in a study done by Tyndale-Boscoe, et al. (2013), there was an overall reversal rate of 17% for signs of use of a handwashing facility and 75% for consistent handwashing with soap and water. Slippage for consistent hand washing with soap and water in Homabay and Kilifi stood at 83% and 67% (Tyndale-Boscoe, et al. 2013). Thus, this study recorded much lower reversal rate on consistent hand washing with soap as compared to previous studies.

Training on hand washing with soap, water treatment, preparing food in a hygienic way and proper storage and solid wastes disposal are standard parts of sanitation program (Magala, & Roberts, 2009). According to Lilje, et al. (2015) in a study done in Chad, the individual perception to treating water was rated high. Respondents thought positively about the issues of water treatment and did not perceive it to be taking much effort, time or cost. This mirrors the findings of this present study, households that treated water were high. Water treatment commodities were available in public health offices and distributed by CHVs at household level during dry seasons, other commodities were offered at health facilities to mothers attending clinics and further, there were chlorine dispensers strategically situated in communal water points.

### Association between social norms and open defecation status

The study found association between a given number of beliefs and expectations and ODF status a reflection of the existence of social norms within Suna West sub county. According to Bicchieri, (2017) for conclusion about the existence of social norms to be arrived at, there is need to be empirical expectation, normative expectation and the belief in the existence of sanctions which by itself is an indication of existence of social norms. Further, this study found that the households that exhibited the social norms were less likely to be ODF.

Results of this study found that household that responded that the health of the family motivates them to be ODF were less likely to be ODF. This is in spite of previous studies like that done by UNICEF (2014), that reported that the most prominent motivator towards ODF status was concern for the health of the family. Households believed that stopping open defecation resulted in reduction in diarrheal diseases thus motivating them to stop open defecation. In another study, Moran, (2017) reports that health, even though may not have been a driver for the initial defecation behaviour change, people do continue to make effort to maintain and use latrine because of their health and that of the family

Further, the study found no association between care for the family, latrine accessibility and ODF status a reflection of no prudential personal normative belief and no association was found in privacy/security offered by latrine and even peer pressure and ODF status. In a previous study, provision for privacy for superstructure, pride and the convenience of using of latrines were found to be important drivers for women in respect to building latrines in Indonesia (Odagiri, et al., 2017). This particular study failed to find an association between shame/disgust and whether it was embarrassing to see people defecate in the open and ODF status while previous studies reported that shame/ disgust motivated households into behaviour change (UNICEF, 2014).

This study found that those who perceived that construction and maintenance materials were expensive (factual belief) were less likely to be ODF, this resonates with studies done previously that found that high cost of building, maintenance and repair of latrines were among the reasons for reversion back to non-ODF status (Mukherjee, 2016). In their study, Bongartz et al. (2016) suggested that though CLTS was a zero-subsidy strategy, there was need for incorporation of sanitation marketing to CLTS to help those who can afford make informed choice even though this could pose challenge of interfering with behaviour change process.

Those that said rewards and incentives motivate them to be ODF were found to be less likely to be ODF. This resonates with the findings of a study done in East Java in which households that received some form of subsidy did not become ODF, it was discovered that subsidy was divisive since it was never enough for all households and thus hampered collective action, also, incentives has been found to have the capacity to corrupt intrinsic motivation (Mukherjee, 2016; Bicchieri, & Noah, 2017). In the study done on sustainability by UNICEF (2014), some of the enablers of sustainability were natural leaders working together and post-ODF follow up by CHVs.

Novotný et al. (2017), in their study, concluded that social norms were important instrumentally as sanitation outcomes depended on the level to which social influences were able to shape the perceptions of benefits or risks on sanitation-related awareness in positive ways. Similarly, Odagiri et al. (2017) found that in addition to economic levels and lack of reliable access to water, weaker social norms were significantly associated with the reversion to open defecation practices. When looked at singly, latrine usage and open defecation were sustained meaning the social sanctions played out well. However, in the other areas of hand washing with soap, provision of privacy and use of squat hole cover, there was significant reversion registered meaning the social sanctions weren’t applied across all the non-negotiable indicators. Suna West sub county had no deep-rooted social norms neither did the CLTS process inculcate new norms to bring about the overall change in sanitation hygiene practices desired and to sustain it after the pressure of certification was off.

## CONCLUSION

There was partial reversion to non-ODF status in households one year after certification of Suna West sub county. This was mainly attributed to 3 major indicators; provision of hand washing facility, squat hole cover and privacy. Moreover, there was sustained ODF status in households that had good sanitation hygiene practices. Social norms were not embedded on the CLTS process, thus failing to create social norms around sanitation and hygiene practices to enhance community collective action towards ODF status sustainability. Therefore, it is important to enhance good hygiene sanitation practices, while instilling social norms to inspire community collective action.

## Data Availability

All data shall be available and will be submitted as part of the manuscript.

## ACKNOWLEDGEMENT

We acknowledge the people of Suna West Sub-County without whom this research would not have been done.

## AUTHORS CONTRIBUTION

N.R.A. conceived the presented idea. N.R.A. developed and performed the study under the supervision of C.O.A. and P.O.O. All authors discussed the results and contributed to the final manuscript.

